# DE NOVO VARIANTS IN THE POLY(RC)-BINDING PROTEIN GENE *PCBP1* CAUSE A NEURODEVELOPMENTAL DISORDER

**DOI:** 10.64898/2025.12.11.25341975

**Authors:** Wallid Deb, Thomas Besnard, Florence Desprez, Benjamin Cogné, Laura Do Souto Ferreira, Virginie Vignard, Sylviane Marouillat, Louis Januel, Svetlana Gorokhova, Tiffany Busa, Victor Morel, Benjamin Dauriat, Vincent Desportes, Anne M Slavotinek, Yu An, Hane Lee, Jessy Hary, Peter Kannu, Taryn B Athey, Ingrid M.B.H van de Laar, Marjon A van Slegtenhorst, Patricia Dickson, Alison M Muir, Rebecca Buchert, Tobias B Haack, Dominic Imort, Sérgio B Sousa, Belinda Xavier, Pedro M Almeida, Mihael Rogac, Borut Peterlin, Sophie Kaspar, Christian Netzer, Hans Zempel, Meghan C Towne, Roger L Ladda, Susan S Sell, Pawel Gawlinski, Xiaofei Song, Wojciech Wiszniewski, Daniel G Calame, Jennifer E Posey, Frederic Ebstein, James R Lupski, Bertrand Isidor, Stéphane Bézieau, Frédéric Laumonnier, Sébastien Küry

**Author notes:** The authors equally contributed to this work (Frédéric Laumonnier, Sébastien Küry). deceased (Roger L Ladda). Corresponding authors: Wallid Deb, Sébastien Küry.

## Abstract

Poly(rC)-binding protein 1 (PCBP1), a splicing factor and key member of the hnRNP E family, was initially characterized for its tumor suppressive properties. More recently, its role in regulating gene expression in the brain and nervous system has attracted growing interest. Through an international multicenter collaboration, we identified 13 *de novo* pathogenic variants in *PCBP1* across 13 unrelated families. All affected individuals exhibited intellectual disability (ID), with autism spectrum disorder (ASD) as a prominent feature. Functional analysis in primary hippocampal mouse neuron cultures demonstrated that *PCBP1* variants impair dendritic arborization, underscoring their deleterious effects. Transcriptomic profiling by RNA sequencing of subjects-derived T cells showed a distinctive signature characterized by significantly increased exon skipping. These results highlight the essential role of PCBP1 in neurogenesis and neuritogenesis, revealing the impact of loss-of-function variants in cells harboring *PCBP1* pathogenic variants, thereby confirming the link between splicing defects and neurodevelopmental disorders. Collectively, our findings demonstrate the critical role of *PCBP1* in neurodevelopment, reaffirming the importance of splicing regulation in mammalian neurodevelopment.

## INTRODUCTION

In eukaryotes, protein diversity is significantly increased through alternative splicing, a critical mechanism in cell physiology by which most messenger RNA (mRNA) precursors (pre-mRNAs) are modified under the action of the splicing machinery. Key components of alternative splicing are RNA-binding proteins (RBPs) ^1^, which in humans are encoded by over 1,500 genes and account for up to 20% of the proteome ^2^. The ubiquitous expression of most RBPs highlights their essential role in the normal processes of human development, particularly in cell differentiation, specification and identity ^2,3^. The two main classes of RBPs are heterogeneous nuclear ribonucleoproteins (hnRNPs) and serine/arginine-rich (SR) proteins, which play a key role in post-transcriptional regulation of gene expression ^4^. By binding to pre-mRNAs and RNAs, RBPs facilitate their assembly with proteins into ribonucleoprotein (RNP) complexes, the best-known example being spliceosomes. They thus contribute to the control of RNA maturation, stability, transport and degradation ^1,2,5^.

In short, alternative splicing is a highly complex mechanism involving a multitude of genes. This complexity, which provides the genetic and protein plasticity essential to cell physiology, also makes alternative splicing highly vulnerable, and up to one in three morbid alleles induces splicing anomalies ^6^. Notably, genetic alterations may disrupt the functions of RBPs in multiple ways. Variants occurring within RBP genes may perturb the formation or localization of RNP complexes through impairment of protein interactions, signal transduction, catalytic activity, or RNA binding ^7^. By contrast, variants located in target RNA may affect their RBP-binding sites, modulating their fate or inducing toxicity ^7^. These various pathological mechanisms are involved in numerous human genetic disorders, ranging from cancers to muscular atrophies and neurological disorders ^7,8^. According to the Online Mendelian Inheritance in Man (OMIM) database, 157 RBPs are currently linked to 221 Mendelian diseases ^7^.

In cancers, most of the RBPs are overexpressed and display oncogenic properties. One of the few RBPs considered as a tumor suppressor is PCBP1 (poly(rC)-binding protein 1) ^9,10^, a member of the poly(rC)-binding protein (PCBP) subgroup of the hnRNP family. Like other PCBP members, PCBP1 is characterized by three conserved KH domains (hnRNP homology domain) which specifically bind to cytosine-rich polypyrimidine motifs ^9–13^, particularly those located in regions 5’ to exonic sequences. This property gives it a special role in modulating the splicing of cassette exons ^13,14^. Among the four known PCBP proteins in humans, PCBP1 and PCBP2 appear to exhibit the highest expression levels and demonstrate both overlapping and exclusive functional roles in mammals ^13,15^. Their distinct roles in several cellular processes have for instance been documented in tumorigenesis ^9^ or cell apoptosis following oxidative stress ^16^. Since PCBP1 and PCBP2 proteins share 83% sequence homology–*the highest among PCBPs*–the non-homologous residues could play a part in their unique tasks ^9,16^.

The high amino acid similarity between these two main PCBP members lies in the origin of PCBP1, encoded by the intronless gene *PCBP*1 (MIM: 601209; MANE select transcript NM_006196.4/ENST00000303577.7), which likely arose from the retrotransposition of a fully processed mRNA of the *PCBP2* gene before mammal radiation ^11,12^. PCBP1 protein sequence is highly conserved, with exact amino-acid residue identity between human (*Homo sapiens*), chimpanzee (*Pan troglodytes*), rhesus macaque (*Macaca mulatta*) and mouse (*Mus musculus*), emphasizing the importance of RNA splicing in molecular diversity, driving the recent evolutionary adaptation of mammals following radiation, alongside genomic recombination and rearrangements ^17–19^. Beyond its evolutionary significance, PCBP1 is abundant in numerous tissues and cell types, as a multifunctional protein regulating not only iron homeostasis and mitochondrial stability as an iron chaperone, but also multiple transcriptional and translational processes through its ability to bind both DNAs and RNAs^20–22^.

Molecular exploration of affected tissues and *in vitro* cell models have hitherto linked *PCBP1* dysfunction to multiple forms of cancer and neurodegenerative diseases such as amyotrophic lateral sclerosis, Huntington disease and Parkinson disease ^3,10,23^. Here, we report constitutional *de novo PCBP1* heterozygous variants found in a series of 13 individuals with developmental delay. We observed significant splicing alterations in cells derived from affected subjects, defining a distinctive transcriptomic signature associated with *PCBP1* deleterious variants. Using a mammalian cell model derived from mouse fetal brains, we demonstrated the impact of such variants on neuronal morphology and regulation of neurodevelopmental processes.

## PATIENTS AND METHODS

### Subjects and genetic analysis

Affected individuals participating in this study were identified in 13 centers in Europe and the United States of America. A candidate variant in *PCBP1* was found in each affected participant during a molecular diagnosis investigation for a neurodevelopmental disorder, primarily characterized by global developmental delay with or without intellectual disability. All variants were identified through routine genetic testing, using pangenomic or gene panel sequencing. The investigating centers were: Nantes University Hospital (Nantes, France); the Baylor Genetics (BG) Laboratories (Houston, TX, USA); the Stollery Hospital (Edmonton, Alberta, Canada); Lyon University Hospital (Lyon, France), the UCSF Benioff Children’s Hospital (San Francisco, USA); the Institute of Human Genetics, University Hospital Cologne (Cologne, Germany); Penn State Health Children’s Hospital *via* Ambry Genetics (Hershey, USA); University Medical Center Ljubljana (Ljubljana, Slovenia); Timone Hospital, APHM (Marseille, France); Washington University in St. Louis *via* the GeneDX laboratory (St Louis, USA); Coimbra University Hospital (Coimbra, Portugal), the Institute of Medical Genetics and Applied Genomics (Tübingen, Germany), and Erasmus Medical Center (Rotterdam, The Netherlands). The connection between the different centers has been facilitated by the web-based tool GeneMatcher ^24^. Written informed consent has been obtained for all affected individuals and relatives. The study has been approved by the CHU de Nantes ethics committee (Advisory Committee on Information Processing in Research; number CCTIRS: 14.556).

### Primary neuronal cultures

All mouse experiments were performed at the University of Tours/INSERM, according to protocols approved by the French Ministry of research (Project authorization number 01456.03 and HC2021-3). Brain tissues were dissected from embryonic day 17.5 C57BL/6J WT mouse embryos (Janvier Labs) in cold DPBS containing 1% of penicillin-streptomycin (ThermoFisher Scientific). The hippocampi were incubated with papain (10 U/mL; Worthington) for 22 min at 37°C, then mechanically dissociated in DMEM:F12 (Gibco, 31331093) with 10% FBS (Eurobio, CVFSVF06-01) and centrifugated at 1100 rpm for 3 min. The cell pellet was resuspended in Neurobasal (Gibco, 21103049) supplemented with B27 (Gibco, 17504044) and GlutaMAX (Gibco,35050061). Dissociated neurons were plated at a density of 450 cells/mm^2^ in 24-well plates on glass coverslips coated with Poly-D-lysine (Merck) and laminin (4 µg/mL; Invitrogen, 23017-015). The cultures were maintained in Neurobasal/B27/GlutaMAX, with half of the medium changed twice a week.

### Transfection

On day 11 of *in vitro* culture, hippocampal neurons were co-transfected with the pCAG-GFP plasmid along with either wild-type or mutant forms of PCBP1-HA constructs, using Lipofectamine 2000 (Thermo Fisher Scientific, 11668500) at a ratio of 0.5 µg of DNA per 1 µL of Lipofectamine 2000 per well.

### Immunochemistry

Neurons were fixed 3 days after transfection, on the 14th day *in vitro*, using a solution of 4% paraformaldehyde - 4% Sucrose. They were incubated for 20 min at room temperature with a fixating solution, washed, and conserved in PBS at 4°C until immunocytochemistry preparation. Fixed neurons were blocked and permeabilized with blocking buffer containing 10% donkey serum / 0.2% Triton X-100 in DPBS for 1 hour, washed in DPBS, and incubated 1 hour with the rat anti-HA High Affinity monoclonal primary antibody (1:200 in blocking buffer; Cat# 11867423001, Roche). After several washes in DPBS, cells were incubated for 45 minutes with the following secondary antibodies diluted in blocking buffer: Alexa Fluor 568-conjugated goat anti-rat IgG (1:500, Cat#A11077, ThermoFisher) or Alexa Fluor 594-conjugated goat anti-rat IgG (1:500; Cat# A11007, ThermoFisher). After three 5-minute washes in DPBS, the stained neurons were mounted in ProLongTM Diamond Antifade reagent (Cat# P36391, 321 Invitrogen).

### Image and statistical analyses

Cellular imaging was performed using a Leica SP-8 laser-scanning confocal microscope and Leica Application Suite X (LAS X) software. Sequential acquisitions were made, and high-resolution z-stack images of cells were captured with either a 40X or 63X objective, with an optical section separation (z interval) of 0.2 µm for the images of the dendritic spines (40X objective, z interval 0.3 µm for the complete neuron). Morphometric and dendritic spine analyses were done with the ImageJ software. Sholl cross-analysis was performed by counting the number of dendrites intersecting concentric spherical radii at 20 µm intervals. Spine density was quantified by calculating the number of spines per dendritic segment and normalized to 10 µm of dendritic length. The ratio of mature/immature spines was determined using morphological criteria: mature spines (mushroom, stubby or branched) had a head diameter ≥0.6 µm, while immature spines (thin and filopodia) had a head diameter <0.6 µm. At least three segments from different dendrites were analyzed for each cell.

Statistical analysis and charts were generated using GraphPad Prism software (La Jolla, CA, USA). Normality of the data was assessed using the Shapiro–Wilk normality test. For neuronal morphological comparison, data were analyzed using two-way ANOVA followed by Sidak’s or Dunnett’s multiple comparisons tests. For spine density analyses, Tukey’s multiple comparisons tests were applied. A Kruskal–Wallis test was done to evaluate the ratio of mature and immature spines. A p-value < 0.05 was considered significant.

### RNA-Sequencing, expression and splicing analysis

RNA-seq experiments were conducted following a similar protocol and workflow as previously described ^25,26^. PBMCs from subjects 1, 5, and 9 were isolated from 2-4 mL of EDTA-treated blood and cultured in lymphocyte-stimulating medium for 48–72 h. RNA was extracted and stranded RNA-Seq libraries prepared from 100 ng total RNA using the SureSelect XT-HS2 kit (Human All Exon V8 capture probes; Ref: G9774C), followed by sequencing on an Illumina NextSeq 550 to obtain ∼25–30 million paired-end reads per sample. Reads were aligned to GRCh38 with STAR v2.7.11a. Quality control was performed with FastQC v0.11.3 and Fastp v0.23.4. Cell type composition was estimated using CIBERSORTx v1.0 with the LM22 signature matrix. For splicing analysis, the subjects carrying loss-of-function variants in *PCBP1* were compared to 49 controls (probands without *PCBP1* variants). rMATS-turbo v4.3.0 was used to detect alternative splicing events with the following parameters: -t paired –anchorLength 1 –libType fr-firststrand –novelSS –variable-read-length –allow-clipping. Python scripts were used to filter rmats output files with the following filters: mean coverage >10, false discovery rate (FDR) < 0.01, |deltaPSI| > 0.10. PCA was performed using the sklearn Python library using PSI values from significant alternatively spliced exons, keeping only (1) the most significant call when several were called impacting the same exon and (2) events affecting genes with approved HGNC symbols. For expression analysis, OUTRIDER (Outlier in RNA-Seq Finder) algorithm was used to detect aberrantly expressed genes as described in literature ^27^. Genes deemed significant outliers were subsequently annotated using the Ensembl database, PanelApp disease panels, pLI scores, and Human Phenotype Ontology (HPO) terms. A significance threshold of p-value < 0.05 was applied for the cohort of 304 samples.

### cDNA targeted Sanger sequencing

To assess the potential involvement of nonsense-mediated mRNA decay (NMD) in our findings, specific primers were designed to amplify by PCR the regions encompassing the *PCBP1* variants identified in subjects 1, 5, and 9 (RefSeq transcript NM_006196.4; Ensembl transcript ENST00000303577.7; MANE Select). Total RNA was extracted from cultured T lymphocytes treated with puromycin (1 mg/mL; Invivogen) for 4 hours, as well as from untreated T cells of the same subjects. Automated Sanger sequencing was then performed.

## RESULTS

### De novo variants in PCBP1 either result in protein truncation or impact a KH domain

Twelve *de novo* single nucleotide variants (SNVs) and one *de novo* multi-nucleotide variant (MNV) were identified in the coding sequence of *PCBP1* (transcript RefSeq NM_006196.4/Ensembl ENST00000303577.7 MANE Select): ten led to a premature termination codon (PTC), including subject 13’s MNV (c.940A>T and c.952_963del, on the same allele), and three were missense SNVs, as detailed in **Figure 1A–B**.

**Figure 1:**
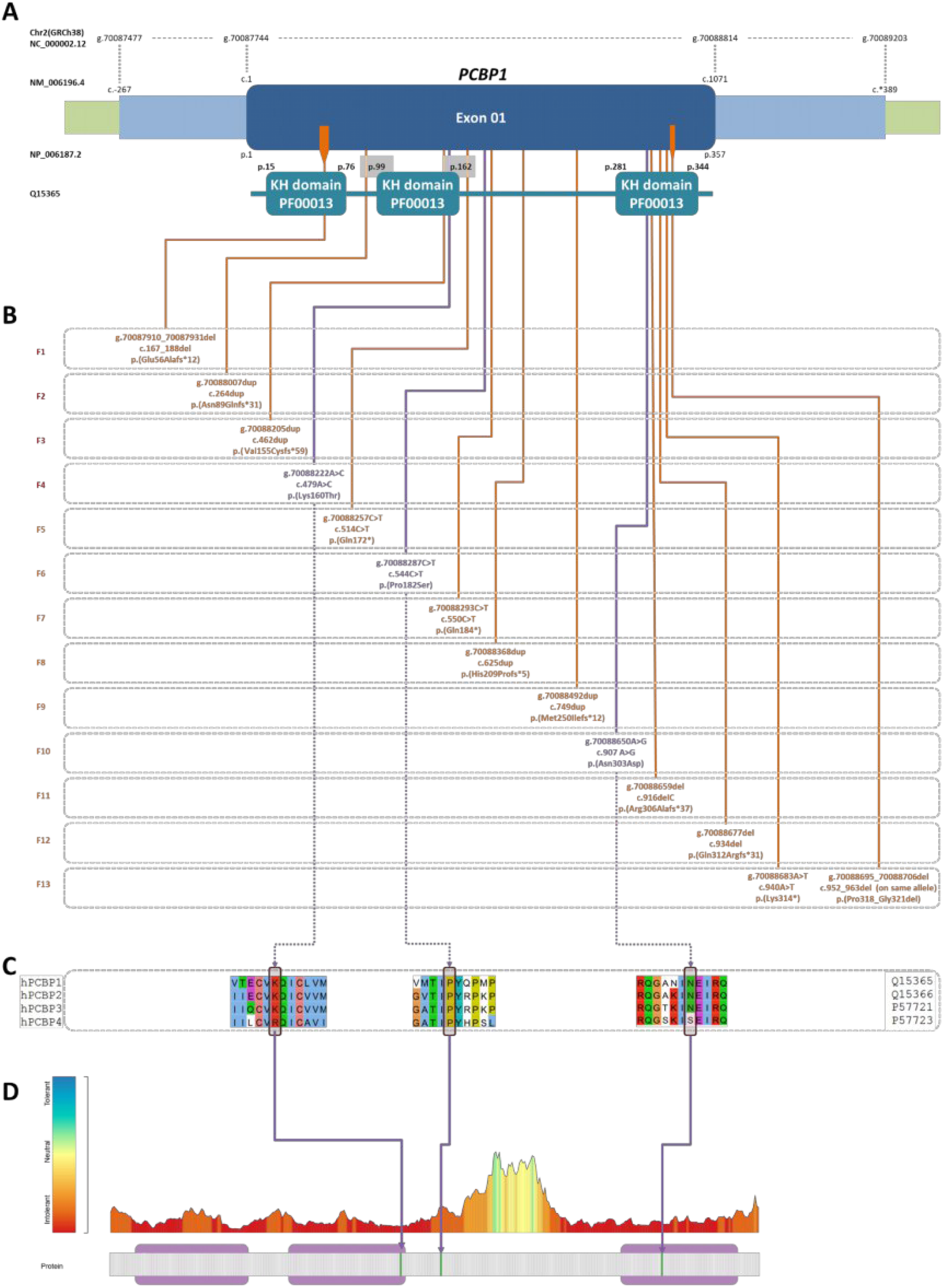
Schematic representation of *PCBP1* and candidate de novo variants. A- Gene and protein domain structure of *PCBP1*. The intronless gene *PCBP1* encodes a protein characterized by three hnRNP K-homology (KH) domains (PF00013), critical for RNA binding. The positions of the identified variants are mapped onto the gene and protein structures. B- PCBP1 variants identified in patients, where each row corresponds to a distinct family (F1–F10). Variants are described using HGVS nomenclature for the genomic (g.), cDNA (c.), and protein (p.) positions. Truncating (orange) and missense (purple) variants are shown with their respective amino acid changes. C-Multiple sequence alignments of human PCBP paralogs (PCBP1-4) illustrate the high conservation of amino acid residues impacted by the three de novo missense variants among the protein family (similar conservation across species too, from human to mouse-not shown). D- Protein sequence constraint and tolerance, Metadome v1.6. The conservation plot (upper panel with color gradient) represents sequence conservation across homologous proteins, with colors ranging from blue (low conservation – high tolerance to variation) to red (high conservation – low tolerance to variation). PCBP1 variants affect highly intolerant residues on the conservation plot, with KH domains highlighted in pink

All the variants were absent in the gnomAD v4.1 control population database. Gene constraint metrics (gnomAD v4) predicted intolerance of *PCBP1* both to protein-truncating variations (pLI=1 with o/e=0) and missense variations (missense Z-score = 4.09). Missense variants p.(Lys160Thr) and p.(Asn303Asp) are respectively located in the second and third KH domains. The p.(Pro182Ser) missense variant lies close to the second KH domain. All amino acid residues impacted by the missense variants are highly conserved among human PCBP proteins and classified as intolerant according to Metadome data (GRCh38, v1.6; **Figure 1C–D**), which is in line with the nucleotide conservation at these positions, with the aforementioned sequence variants all displaying a CADD Phred score above 20. Detailed bioinformatics interpretation of pathogenicity by the MobiDetails annotation platform ^28^ using ACMG criteria ^29^ are available for all of these variants following the individual variant links provided in **Table S1**.

Given the identification of *de novo* heterozygous truncating and highly damaging missense variants in functional domains, we hypothesized that the disruption of *PCBP1* protein product would result in a loss of function.

### Individuals harboring a de novo *PCBP1* variant show intellectual disability, global developmental delay, autism and ophthalmological anomalies

A clinical summary of the main features is provided in **Table 1**. No family history of neurodevelopmental disorders was reported in twelve of the thirteen unrelated and non-consanguineous families. Subject 4 had a paternal cousin with Asperger syndrome and a maternal cousin with autism, though no further details were available. Sporadic presentation with *de novo* variants was therefore considered the expected mode of onset.

**Table 1.** Clinical recap of main features in individuals carrying a *PCBP1* variant (n=13)

|  |  |  |
| --- | --- | --- |
| Gender | F=4 | M=9 |
| Age | 1y-21yo |  |
| Growth |  |  |
| Low birth weight (<10p) | 2 |  |
| Microcephaly (≤-2DS) | 3 | |
| Macrocephaly (≥2SD) | 0 | |
| Short stature (≤-2SD) | 2 | |
| Neurobehavioral |  |  |
| Intellectual disability (/assessed) | 10/10 |  |
| Borderline/mild | 5 |  |
| Moderate | 2 |  |
| Severe/profound | 3 |  |
| Developmental delay | 13 |  |
| Motor delay | 11 |  |
| Hypotonia | 9/12 |  |
| Speech delay | 13 |  |
| Abnormal behavior | 10/12 |  |
| Autism/ASD | 8 |  |
| ADHD | 3 |  |
| Seizures | 3 |  |
| Miscellaneous |  |  |
| Ophtalmological | 6 |  |
| Cardiac malformation | 3 |  |
| Urogenital | 2 |  |

All individuals from the cohort, nine males and four females, exhibited neurodevelopmental delay (13/13). They all presented a variable degree of intellectual disability (ID), ranging from mild (n=5) to severe (n=3). Speech delay was found in all subjects (n=13), regardless of the severity of ID. When the information was available, motor delay was also frequent, though not constant (n=11), with delayed walking, starting typically after 20 months. Hypotonia was noted in 9/12 assessed individuals (severe in n=2) mostly in subjects with subsequent severe motor delay.

Neurobehavioral phenotyping revealed autism or autism spectrum disorder (ASD) as a prominent feature in 9 out of 12 assessed individuals, none of whom had been initially addressed and evaluated for ASD specifically.

Ophthalmological anomalies were frequent (n=6), including optic nerve hypoplasia in two subjects, refractive anomalies in three subjects, and strabismus in two subjects. One patient showed a retinal involvement (suspected dystrophy with pigmentary shift).

Regarding congenital malformations, we observed congenital heart defects in three subjects, and urogenital malformations (pyelectasis and asymmetric kidneys) along with cryptorchidism and inguinal hernias in two subjects.

Although variable dysmorphic features were observed among affected individuals, no facial gestalt or shared pattern of recognizable facial features or dysmorphism could be defined. A detailed list of specific facial traits and detailed clinical features is provided in **Table S2**.

### Functional analysis of *PCBP1* variants in mature primary neuronal cultures reveal impaired PCBP1 dendritic sorting or arborization processes

To directly assess the impact that *PCBP1* variants would cause on neuronal development and synapse formation and maturation, we used primary neuronal cultures derived from the dissection of mouse embryonic brains (at the 17^th^ day of gestation), specifically dissociating the hippocampal structure. Interestingly, PCBP1 and PCBP2 proteins have been detected in the postsynaptic density proteome of both human and mouse brain, suggesting a particular function within the synaptic environment ^30,31^.

At day 11 of neuronal culture, after a significant proportion of neurons have reached maturity and developed synaptic structures, we conducted co-transfection experiments using pCAG-GFP (GFP labeling of the entire neuron from soma to dendritic spines) and HA-tagged PCBP1 (PCBP1-HA) wild type (WT) or variant expression plasmids. The variant plasmids were generated by in-site directed mutagenesis applied to the PCBP1-WT plasmid (see PATIENTS AND METHODS). All constructs were validated by Sanger sequencing analyses. The transfected neuronal cultures were fixed 72 hours post-transfection, immuno-stained with primary anti-HA antibodies to detect PCBP1-HA proteins, and visualized by confocal microscopy.

We first observed that overexpression of PCBP1-HA WT led to a significant increase in dendritic arborization when compared to control neurons (i.e., neurons transfected with pCAG-GFP alone), suggesting that PCBP1 would promote dendritic development (**Figure 2 A-B**). Upon overexpression of PCBP1-HA variants, we detected diverse effects on PCBP1 subcellular localization or dendritic arborization. Most notably, the p.(Glu56Alafs*12) PCBP1 protein displayed a markedly reduced signal in dendrites compared to the PCBP1-WT protein (**Figure 2C**), suggesting that its sorting and/or trafficking to dendrites may be altered. In contrast, the four other variants, including the truncating p.(Gln184*) variant, did not show any alteration in the dendritic localization of PCBP1 (**Figure 2C**).

**Figure 2.**
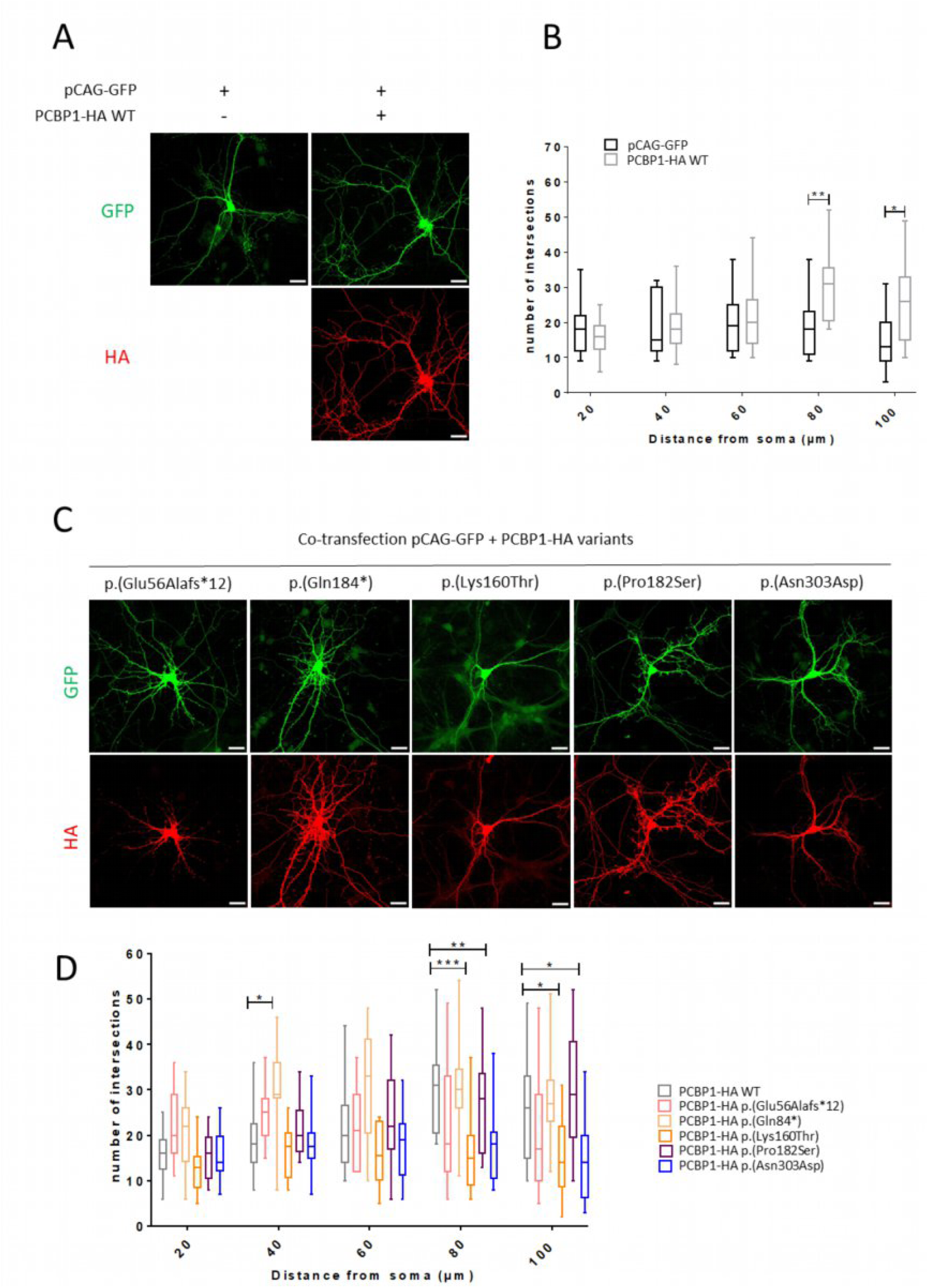
Functional analysis of PCBP1 protein variants in primary cultures of mouse hippocampal neurons. A- Representative images from confocal microscopy showing mature primary neurons co-transfected at 11 days in vitro (DIV) with PCBP1-HA wild type (WT) protein. Co-transfection with pCAG-GFP (green fluorescent protein) was performed to visualize the entire cell. Neurons were stained at DIV 13 for the assessment of subcellular localization of PCBP1 and of dendritic arborization by Sholl analysis. The anti-HA antibody revealed the subcellular localization of PCBP1-HA WT or variants forms (red fluorescence). B- Sholl analysis of pCAG-GFP or PCBP1-WT transfected neurons. Overexpression of PCBP1-WT increased arborization from 80 µm of the soma compared to control neurons transfected with pCAG-GFP. Data are presented as mean±SEM, n = 11 pCAG-GFP neurons and 13 WT-PCBP1 neurons from 3 independent transfections. Two-way analysis of variance with Sidak’s multiple comparisons tests (20 µm, p>0,9999; 40 µm, p>0,9999; 60 µm, p>0,9999; 80 µm, **p<0.01; 100 µm, * p<0.05). C- Representative images from confocal microscopy showing mature primary neurons co-transfected at 11 days in vitro (DIV) with p.(Glu56Alafs*12), p.(Gln184*), p.(Lys160Thr), p.(Pro182Ser), and p.(Asn303Asp) PCBP1-HA variants. Co-transfection with pCAG-GFP (green fluorescent protein) was performed to visualize the entire cell. Neurons were stained at DIV 13 for the assessment of subcellular localization of PCBP1 and of dendritic arborization by Sholl analysis. The anti-HA antibody revealed the subcellular localization of PCBP1-HA variants (red fluorescence). The p.(Glu56Alafs*12) variant impaired the dendritic localization of PCBP1. X40 objective, scale bar : 30 µm. D- The variants p.(Lys160Thr), p.(Asn303Asp) lead to a loss of effect of PCBP1-WT overexpression on dendritic arborization complexity. Data are presented as mean ±SEM, n = 13 PCBP1-WT, 11 p.(Glu56Alafs*12), 12 p.(Gln184*), 14 p.(Lys160Thr), 13 p.(Pro182Ser) and 12 p.(Asn303Asp) neurons from 3 independent transfections. Two-way analysis of variance with Dunnett’s multiple comparisons tests (20 µm, p>0,9999 ; 40 µm, *p<0.05 ; 60 µm, p>0,9999 ; 80 µm, **p<0.01 and ***p<0.001; 100 µm, *p<0.05).

Regarding the dendritic arborization analyses, our experiments revealed variable consequences: (1) neurons overexpressing the p.(Gln184*) PCBP1-HA proteins exhibited a significantly increased number of dendritic intersections compared to PCBP1-HA WT neurons at 40 µm from the soma; (2) neurons overexpressing the p.(Lys160Thr) and p.(Asn303Asp) variants displayed significantly decreased dendritic arborization compared to PCBP1-HA WT neurons when measured beyond 60 µm from the soma (80 and 100 µm) (**Figure 2C–D**); (3) overexpression of the p.(Pro182Ser) variant did not induce any significant change. Our data thus suggest that overexpression of the variants, with the exception of p.(Pro182Ser), disrupt the physiological processes regulating the maintenance of neuronal dendritic arborization in mature neurons.

We next investigated the impact of PCBP1 variants on synaptic density and morphology. Although we previously established that the p.(Glu56Alafs*12) PCBP1 protein was not properly expressed in dendrites and at the dendritic spines, our analyses did not reveal any significant variation in the number or morphology of dendritic spines in mature neurons overexpressing pCAG-GFP alone, pCAG-GFP and PCBP1-HA WT, or pCAG-GFP and the candidate variants (**supplementary Figure S1**).

Our findings highlight three key results: (1) PCBP1 plays a crucial role in neuronal development, as the overexpression of WT-PCBP1 promotes dendritic arborization; (2) PCBP1 variants impair dendritic growth in different ways, with some enhancing and others reducing arborization; and (3) despite the altered dendritic localization observed for the p.(Glu56Alafs*12) variant, none of the tested variants displayed a significantly affected synaptic density or morphology when overexpressed. These results suggest that PCBP1 contributes to neuronal maturation primarily by regulating dendritic development in mammals, while its role in synaptic structure could not be observed through this work, and may be more subtle or context-dependent.

### RNA-seq in subject-derived T cells shows a splicing signature characterized by an increase in exon skipping

Blood samples were obtained for subjects 1, 5, and 9 carrying the p.(Glu56Alafs*12), p.(Gln172*) and p.(Met250Ilefs*12) *PCBP1* variants, respectively. Isolation and culture of T cells, followed by RNA-sequencing (as detailed in PATIENTS AND METHODS), allowed us to evaluate the transcriptome of patient-derived cells carrying a heterozygous loss-of-function variant in *PCBP1*.

Using the OUTRIDER algorithm to detect aberrant transcript expression, we detected a loss of *PCBP1* transcript expression in cells carrying the p.(Glu56Alafs*12) variant, with a -0.7 log2 Fold change (0.6-fold change; p-value = 1.24x10^-15^, adjusted p-value = 5.36x10^-10^), in line with an almost complete monoallelic expression of the wild-type allele with haploinsufficiency, which has been confirmed by analyzing the aligned RNA-Sequencing files. This decreased expression was not seen for the other two variants p.(Gln172*) and p.(Met250Ilefs*12), which was expected since *PCBP1*, as a monoexonic gene, is in theory not regulated by the same Nonsense-Mediated Decay (NMD) rules that apply to premature termination codons in multi-exon genes ^32^.

Then, we sought to explore the potential splicing defects in subjects carrying a *PCBP1* loss-of-function variant. To achieve this, we used rMATS-turbo ^33^ and identified significant aberrant splicing events when comparing cells from subject 1, 5, and 9 to 49 controls (individuals with NDDs, not carrying a *PCBP1* candidate variant). For each category of splicing defects, we identified a number of significant events (FDR < 0.01, |ΔPSI| > 0.1): 116 for exon skipping (ES), 39 alternative 5′ splice sites (A5SS), 50 alternative 3′ splice sites (A3SS) and 47 intron retention (RI) in total (Fig 3A). We then extracted the percent spliced in (PSI) values of these events, and performed a Principal Component Analysis (PCA) on PSI values of significant events. With this approach, the most discriminant effect was from the exon skipping events, with a clear and distinct cluster for the subjects in comparison with the control group as shown on **Figure 3B**. Other PCA plots for A5SS, A3SS and RI are presented in **supplementary Figure S2.** When focusing on the PSI value (<0 for an increased exon skipping in subjects; >0 in favor of an increased exon skipping in controls), we see an almost exclusive representation of increased exon skipping events in subjects carrying *PCBP1* loss-of-function variants, as shown in **Figure 3C**. We can also observe that the exon skipping increase seen in *PCBP1* variant cells affect already existing skipping events in controls, as illustrated in **Figure 3D**.

**Figure 3.**
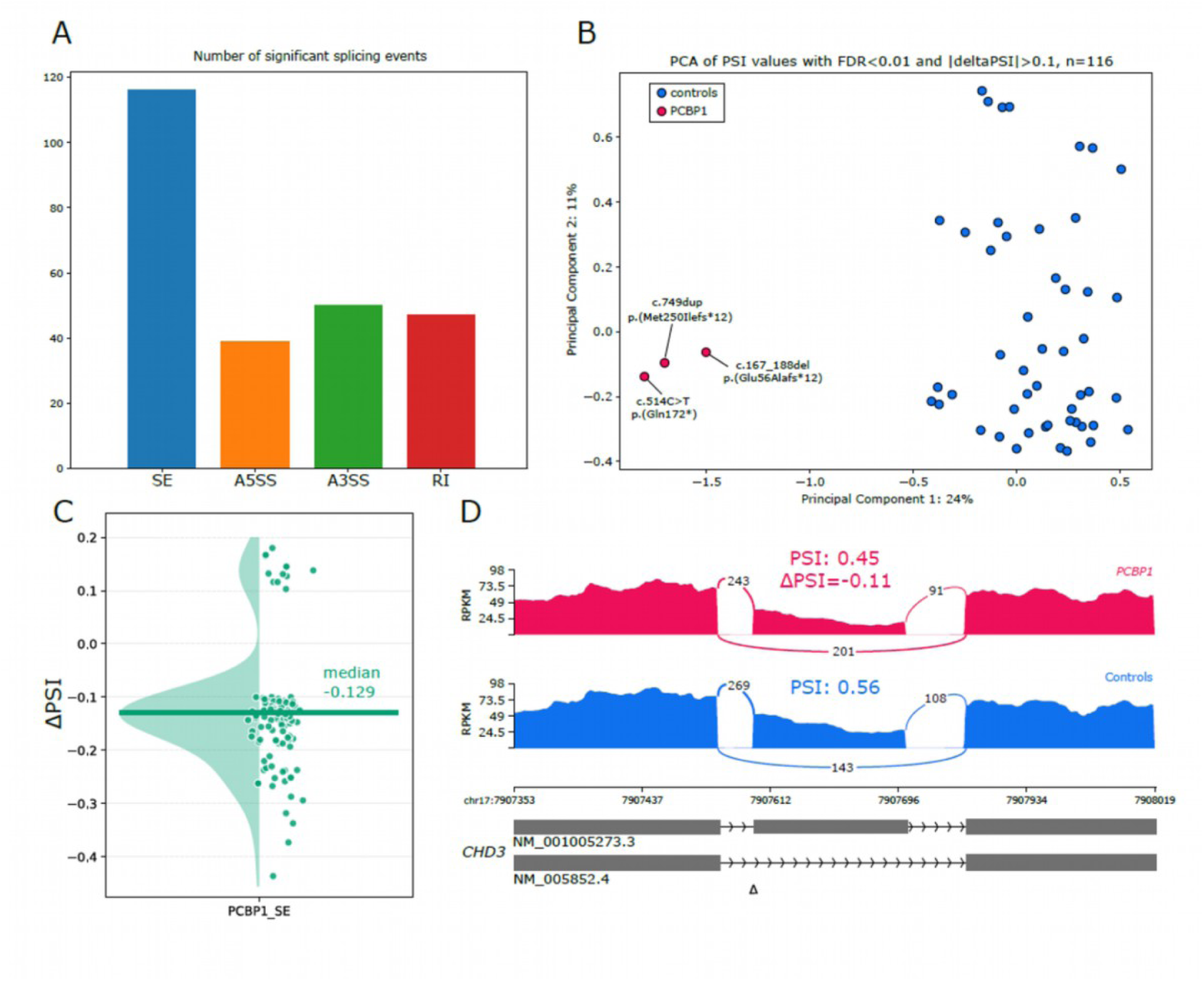
RNA-Sequencing identifies an exon-skipping signature in patient-derived cells harboring loss-of-function variants in *PCBP1*. A- Number of significant alternative splicing events (|ΔPSI| > 0.1; FDR <0.01) detected using rMATS in 3 subjects with a PCBP1 variant, compared to 49 controls. The four splice event categories are colour-coded: exon skipping (SE, blue; n=116), alternative 5′ splice sites (A5SS,orange; n=39), alternative 3′ splice sites (A3SS, green; n=50), and intron retention (RI, red; n=47). B- Principal component analysis (PCA) of PSI values for the 116 significant ES events, showing a clear separation between 3 patients with a PCBP1 variants (red; subject 1 p.(Glu56Alafs*12); subject 5 p.(Gln172*) and subject 9 p.(Met250Ilefs*12)) and 49 controls (blue). C- Distribution of ΔPSI values for significant events (threshold: |ΔPSI| > 0.1), showing a majority of increased exon skipping (negative PSI value) with a median of -0.129 for all significant events. D – Sashimi plot of a representative physiological exon-skipping event in CHD3, showing reduced exon inclusion in PCBP1 subject cells (upper track, PSI = 0.45) compared to controls (lower track, PSI = 0.56), consistent with increased exon skipping (ΔPSI = −0.11) associated with PCBP1 loss of function.

RNA-Sequencing in subject-derived T cells revealed three key results: (1) the p.(Glu56Alafs*12) appears to have an impact on *PCBP1* transcript expression, with a plausible haploinsufficiency from expression data; (2) a transcriptomic signature discriminating subjects from controls; (3) characterized mainly by abnormally increased exon skipping events in subjects carrying a *PCBP1* variant.

### Potential role of Nonsense-Mediated Decay in patient derived T cells carrying a loss-of-function variant in PCBP1

To assess the unexpected decreased *PCBP1* transcript expression following OUTRIDER RNA-Seq expression results, we amplified *PCBP1* transcripts in subject 1, 5, and 9 harboring the variants c.167_188del, p.(Glu56Alafs*12); c.514C>T, p.(Gln172*) and c.749dup, p.(Met250Ilefs*12), respectively, followed by Sanger sequencing of puromycin-treated and untreated cells. When comparing cDNA sequencing in cells carrying the c.167_188del, p.(Glu56Alafs*12) variant, we could see an increase in the expression of the transcript harboring the 22 bp frameshift deletion, suggesting an impact of puromycin, potentially inhibiting degradation of the *PCBP1* mutant mRNA. This effect could not be replicated in cells carrying the c.514C>T, p.(Gln172*) and c.749dup, p.(Met250Ilefs*12) variants, with a clear heterozygous representation of wild type and variant sequence. The automated Sanger sequencing results are shown in **supplementary Figure S3**.

## DISCUSSION

Our study sheds light on a neurodevelopmental disorder caused by variants in *PCBP1*, a gene encoding a trans-acting splicing protein extensively studied for its role in cancerous and neurologic conditions ^9,10,14,23^. In the *PCBP1*-related NDD that we describe, the identification of primarily *de novo* loss-of-function and highly damaging missense variants suggests that loss of function is the underlying cause of the phenotype observed in affected subjects, as suggested by the results in T cells harboring the c.167_188del, p.(Glu56Alafs*12) variant and the RNA-Sequencing data. This hypothesis is in line with the pathogenic model described for the *NOVA2*-associated NDD, where loss of function, partial in that case, can significantly impair alternative splicing of at least 41 target genes, several of them playing a critical role in neurodevelopment ^34,35^. Notably, haploinsufficiency is also at the origin of the dysregulations observed in carcinogenesis that results from the loss of PCBP1 tumor suppressor functions ^9,14,36^.

In the subjects reported here, *PCBP1* variants are primarily associated with neurodevelopmental delay, frequently accompanied by facial dysmorphism–though not recognizable–and ophthalmological features. Given the ubiquitous expression of *PCBP1* across tissues, a broader clinical spectrum might have been anticipated. Transcriptomic data from GTEX v10 indicate widespread *PCBP1* expression (transcript median levels ranging from 34 to 335 TPM), with relatively uniform distribution across cerebral regions, despite overall lower expression in the brain (34–97 TPM). The Human Protein Atlas similarly confirms a high and broadly distributed abundance of the PCBP1 protein. The predominance of neurodevelopmental manifestations suggests that neuronal cells may be more vulnerable to the loss of PCBP1. This is further supported by the crucial and prevalent role of alternative splicing in the nervous system, more than in any other tissues, which underpins its functional complexity ^3,18,31^.

In fact, multiple studies have shown the essential role of PCBP1 target RNAs in critical processes in the central nervous system, supporting the idea that disruption of *PCPB1*-related splicing might impair physiological regulation and functioning of key neurodevelopmental pathways. For instance, in mice, the µ-opioid receptor (MOR), encoded by *OPRM1* and involved in brain development and homeostasis, can undergo transcriptional regulation mediated by PCBP1 functioning in that case as a DNA-binding protein ^37^. In the human neuroblastoma cell line SH-SY5Y, PCBP1 knock-down (KD) induces significant dysregulation of the proteome, in molecular pathways crucial for physiological neurogenesis, axonogenesis, and neurite morphogenesis, even under a conservative assessment ^21,38^. Considering these examples, along with numerous other studies reporting the effects of PCBP1 in the nervous system, it appears more plausible that *PCBP1*-related neurodevelopmental disorder derives from the dysregulation of a great number of PCBP1 target genes, as shown in PCBP1 loss of function in oncogenesis ^9,14,36^. Our results in mice-derived cellular models are consistent with the published data, showing several variants causing an abnormal dendritic sorting and arborization process. The p.(Pro182Ser) variant, while being a strong candidate, does not appear to impact these processes significantly. Its specific effect may, in part, stem from its location outside a functional KH domain, which needs to be investigated further. Considering the impact of the other variants, this work suggests a role of PCBP1 in processes that have already been linked to neurodevelopmental disorders. Dendritic architecture is indeed a critical determinant of neural networks formation and complexity, and alteration of dendritic morphology, such as increased or decreased dendrite number and branching, is associated with intellectual disability and ASD in humans and in animal models ^39–41^.

RNA-Seq demonstrated that subject-derived cells harboring loss-of-function variants in *PCBP1* exhibit a distinct transcriptomic signature characterized by aberrant splicing events. Notably, most significant findings involved increased exon skipping, consistent with at least partial loss of function caused by deleterious *PCBP1* variants. These splicing abnormalities might critically contribute to the predominantly neurological and developmental phenotype observed in affected individuals, although the downstream impact on molecular actors involved in neurodevelopment remains to be elucidated. In addition, this transcriptomic signature could serve as a valuable tool for future assessment of variants of unknown significance, facilitating improved diagnostics as previously established using the same RNASeq analysis protocol ^25^. A noteworthy observation was the apparent activation of nonsense-mediated decay (NMD) in cells carrying the c.167_188del, p.(Glu56Alafs*12) variant, despite *PCBP1* being a monoexonic gene. While initially unexpected, this could very well be explained by the involvement of Exon Junction Complex-independent NMD (non-EJC-NMD), as described for protein truncating variants located upstream of unusually long or unstructured 3’UTR (exceeding 1kb) ^32,42^. As it occurs, the c.167_188del, p.(Glu56Alafs*12) variant induces a premature termination codon more than 1.2 kb before the end of the newly extended 3’UTR, thereby potentially triggering non-EJC-NMD. Although further investigation is required, the absence of mutant allele in untreated subject 1-derived cells supports the hypothesis of loss of function and haploinsufficiency. However, correlation between phenotype and *PCBP1* expression levels could not be determined in this work and will require additional samples from subjects carrying early PTC variants.

These findings align with studies showing the essential role of RBPs in the nervous system, and how variants in RBPs can disrupt their function during corticogenesis and result in severe neurodevelopmental disorders ^31,43^. For instance, the spliceosome component RBM8A (encoded by the eponymous gene), and the RNA-binding protein FMRP (encoded by *FMR1*) have been associated with developmental disorders affecting the central nervous system: TAR syndrome [MIM: 274000] and Fragile X syndrome [MIM: 300624], respectively ^31^. More recently, the gene for the NOVA alternative splicing regulator 2 (*NOVA2*) ⎯ which possesses three KH domains like PCBP1⎯ previously associated with the paraneoplastic opsoclonus myoclonus ataxia syndrome (POMA), has been shown to cause a severe autosomal dominant neurodevelopmental disorder [MIM: 618859] ^35^. The link between RBPs and neurodevelopmental disorders is increasingly documented, with various pathogenic mechanisms involved: dysregulation of mRNA stability and/or translation (FMRP), localization of target RNA, or dysregulation of alternative splicing (RBM8A, NOVA2) among others^31,34,35^.

Given the immense molecular and cellular diversity and complexity introduced by alternative splicing, it makes sense that its functions brought layers of phenotypic combinations that could have played a part in the massive radiation that occurred between 60 to 120 million years ago, and in the evolution of the mammalian brain as we know it today ^17,18,44^. The recent incorporation of *PCBP1* into the mammalian genome further suggests that PCBP1 and other RBPs may have a key influence on gene expression in traits specific to mammals, one of which being the development of the brain and nervous system. Further research will be necessary to pinpoint the precise impact of PCBP1 on transcript and isoform expression in the human and mammalian cells and brain, and to fully understand its role in brain development and pathology.

## Supporting information

AllSupplementary

## Data Availability

All data produced in the present study are available upon reasonable request to the authors

## Data and code availability

All raw data presented in this paper are available to qualified researchers upon request.

## ACKNOWLEDGMENTS

This work has been carried out within the framework of the FHU GenOMedS thanks to the support of the Health cooperation group of University Hospitals of the Great West (GCS HUGO). This work was supported by the Groupama Foundation.

F.E. acknowledges the I-SITE NExT Junior Talent Chair. S.K., and F.L. received research grants from the Agence Nationale de la Recherche (ANR) for projects ANR-21-CE17-0005 (UPS-NDDecipher) and ANR-22-RAR4-0001-01 (UPS-NDDiag), as well as support to S.K. from la Région des Pays de la Loire for project TN_2021_AAP_UPS-NDDECIPHER_INSERM_154550. EJP RD funding was received from the European Union’s Horizon 2020 research and innovation program under grant agreement N°825575. S.B. received financial support from the University Hospital Center (CHU) of Nantes for the BioTND-UPS biobank (PROG/09/72-03), and S.B., S.K., F.L., and F.E. received funding from Mutuelles AXA for project TND-UPS. J.R.L. and D.G.C. received support from various sources including the US National Institutes of Health (R35 NS105078 and U01 HG011758), Rett Syndrome Research Trust, International Rett Syndrome Foundation, Doris Duke Charitable Foundation, Blue Bird Circle Foundation, and the Muscular Dystrophy Association.

## Author contributions

W.D., T.Besnard, B.C., S.B., F.L., and S.Küry conceived the study and designed the experiments. W.D. and S.Küry supervised subject enrollment. Genomic data analysis was supervised by W.D., T.Besnard, B.C., and S.Küry.

F.D., S.M., and F.L. designed, conducted and analyzed all the functional studies in mouse mature primary neuronal cultures. B.C., L.dS, T.B., S.Küry, and W.D. designed, conducted, and analyzed the RNA-Sequencing data. Clinical data were collected and analyzed by W.D., T.B., B.I., S.B., and S.Küry

B.I, B.D., C.N., H.Z, L.J., S.G., S.Kaspar, T.Busa, V.D., V.M. provided critical blood samples necessary for generating RNA-sequencing data.

A.M.S., A.M.M, B.D., B.I., B.P., B.X, C.N., D.G.C., D.I., F.D., F.E., F.L., H.L., H.Z., I.M.B.H.vL., J.E.P., J.H., J.R.L., L.dS., L.J., M.A.vS., M.C.T., M.R., P.D., P.G., P.K., P.M.A., R.B., R.L.L., S.B.S., S.G., S.Kaspar, S.M., S.S.S., T.B.A., T.Busa, T.H., V.D., V.M., V.V., W.W., X.S., Y.A provided critical material or data to carry out the research.

Funding acquisition was secured by S.Küry, B.C., and S.B.

All authors reviewed the manuscript, figures and data.

## Web resources

Combined Annotation Dependent Depletion (CADD): https://cadd.gs.washington.edu/

GenBank: http://www.ncbi.nlm.nih.gov/genbank/

gnomAD: http://gnomad.broadinstitute.org/

GTEx: https://www.gtexportal.org/home/

Human Protein Atlas: https://www.proteinatlas.org/

Metadome: https://stuart.radboudumc.nl/metadome/faq

Mobidetails: https://mobidetails.chu-montpellier.fr/

OMIM: http://www.omim.org/

UniProt: http://www.uniprot.org/uniprot/

