## Supplementary material for "DE NOVO VARIANTS IN THE POLY(RC)-BINDING PROTEIN GENE *PCBP1* CAUSE A NEURODEVELOPMENTAL DISORDER": AllSupplementary

**Figure S1**. The overexpression of PCBP1 variants does not alter synapse density and maturity processes in neurons

**Figure S2**. PCA based on PSI values in T cells carrying loss of function variants in *PCBP1*.

**Figure S3.** Effect of puromycin treatment on variant *PCBP1* transcripts in subject-derived T cells.

**Table S1**. Molecular characteristics of candidate *de novo* variants in *PCBP1*

**Figure S1. The overexpression of PCBP1 variants does not alter synapse density and maturity processes in neurons**

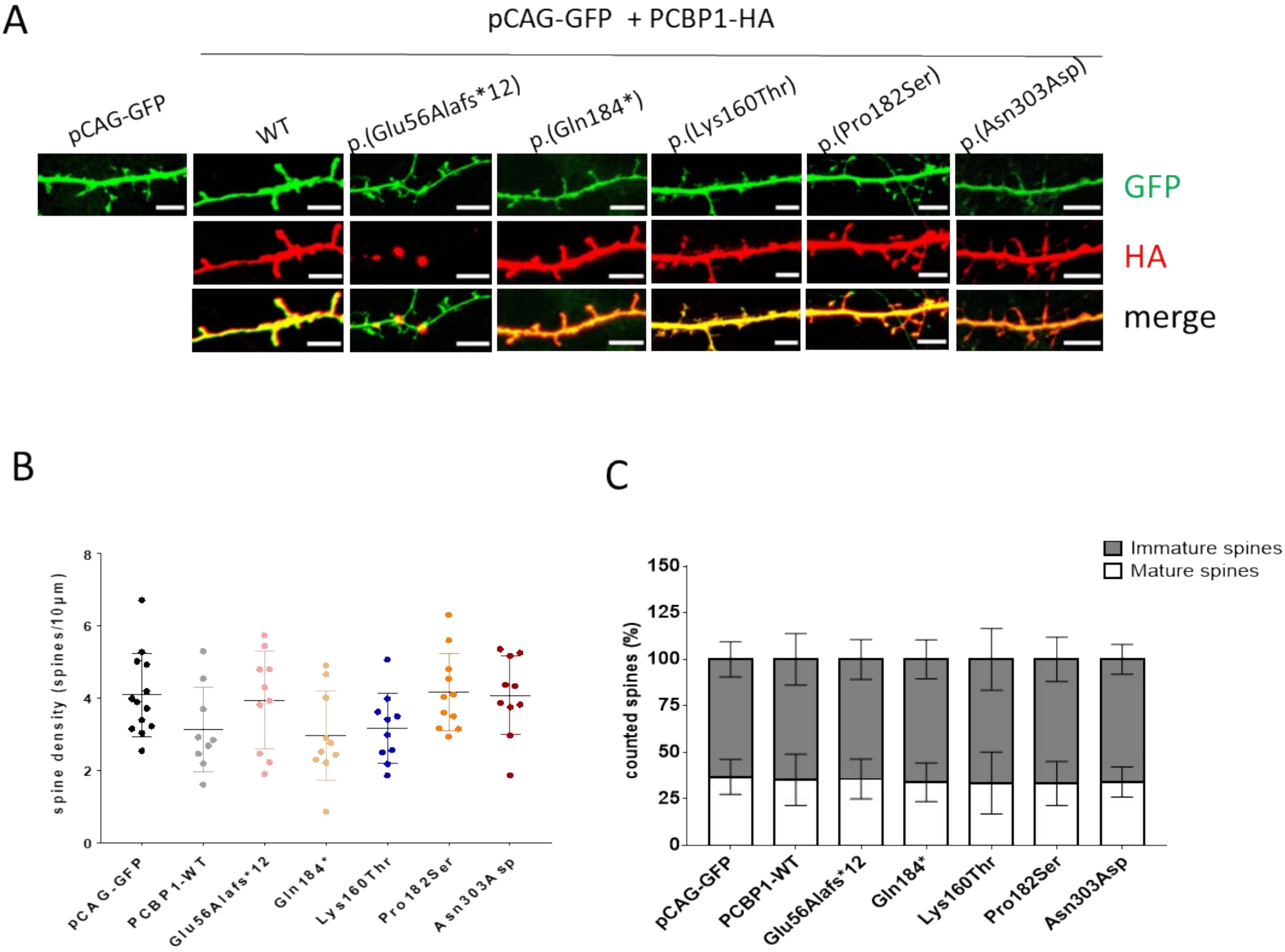

**Figure S1. The overexpression of PCBP1 variants does not alter synapse density and maturity processes in neurons**

**A-** Representative images from confocal microscopy showing dendrite sections and dendritic spines of mature hippocampal neurons. At *day in vitro (DIV)* 11, neurons were co-transfected with pCAG-GFP and wild-type (WT) or mutant forms of PCBP1. Immunocytochemistry was performed using an anti-HA (PCBP1-HA, red). The GFP protein was used to visualize spine morphology. X63 objective, scale bar : 5µm. **B-** Graphical representation of the spine density analysis for each PCBP1 variant. The data are represented as mean± SD, *n* = 13 pCAG-GFP, 9 PCBP1-WT, 10 p.(Glu56Alafs*12), 10 p.(Gln184*), 10 p.(Lys160Thr), 11 p.(Pro182Ser), 10 p.(Asn303Asp) neurons from 3 independent transfection. One-way analysis of variance with Tukey's multiple comparisons (no significant, p>0,9999). **C-** Graphical representation on the analysis of the ratio mature/immature spines for each PCBP1 variant. Data are represented as mean± SD, n= 13 pCAG-GFP, 9 PCBP1-WT, 10 p.(Glu56Alafs*12), 10 p.(Gln184*), 10 p.(Lys160Thr), 11 p.(Pro182Ser), 10 p.(Asn303Asp) neurons from 3 independent transfection. One-way analysis of variance with Kruskal–Wallis Test (no significant, p>0,9999).

**Figure S2. Transcriptomic signature in patient-derived cells harboring loss of function**

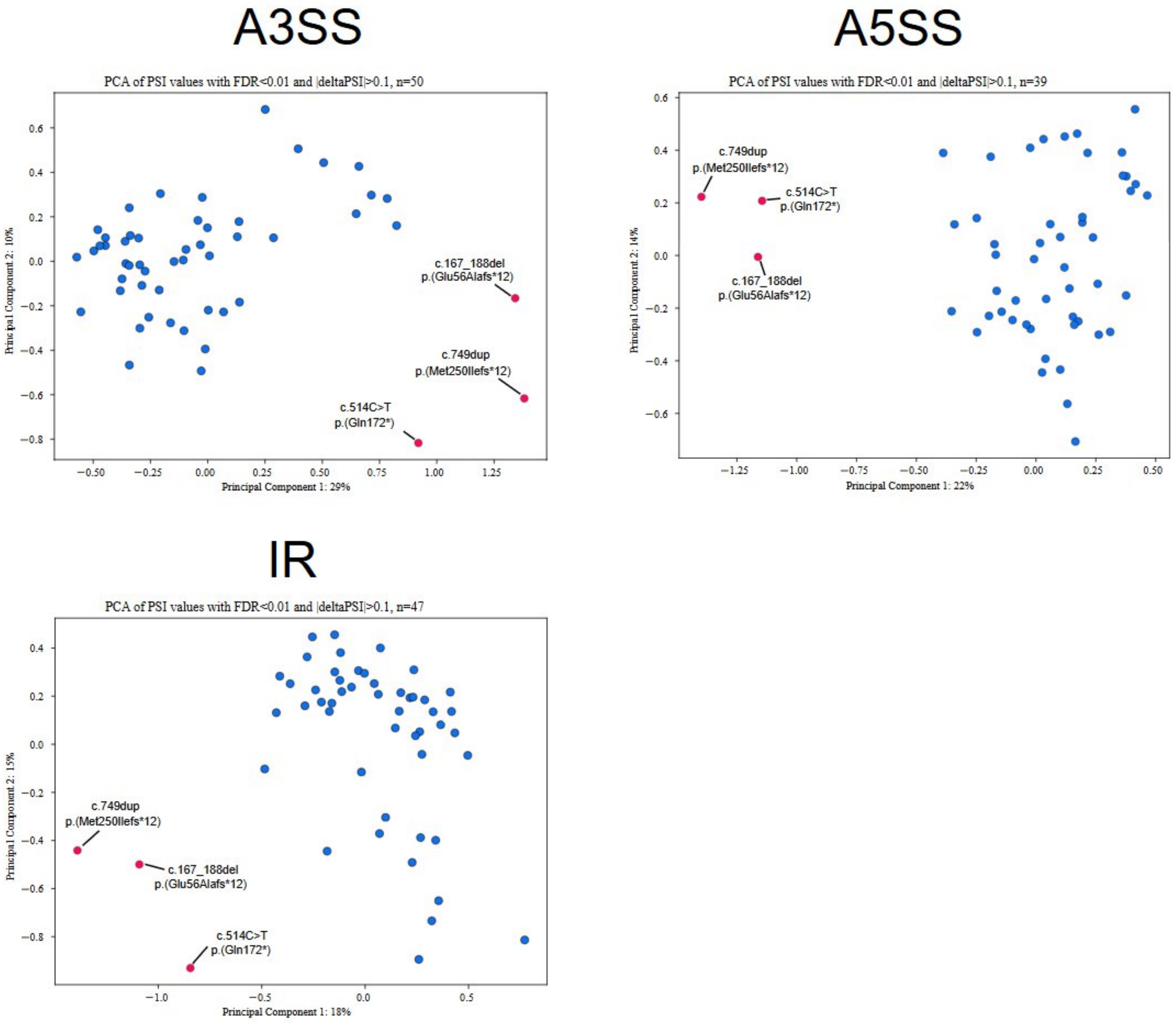


**variants in *PCBP1*.**

**Figure S2. Transcriptomic signature in patient-derived cells harboring loss of function variants in *PCBP1*.**

Principal component analysis (PCA) of PSI values for the significant events, by category: 50 alternative 3′ splice sites events (A3SS, top-left) ; 39 alternative 5′ splice sites events (A5SS, top-right), and 47 intron retention (RI, bottom-left). A clear separation is observed between the 3 subjects with a *PCBP1* variants (red; subject 1 p.(Glu56Alafs*12); subject 5 p.(Gln172*) and subject 9 p.(Met250Ilefs*12)) and the 49 controls (blue).

**Figure S3. Effect of puromycin treatment on variant *PCBP1* transcripts in subject-derived T cells.**

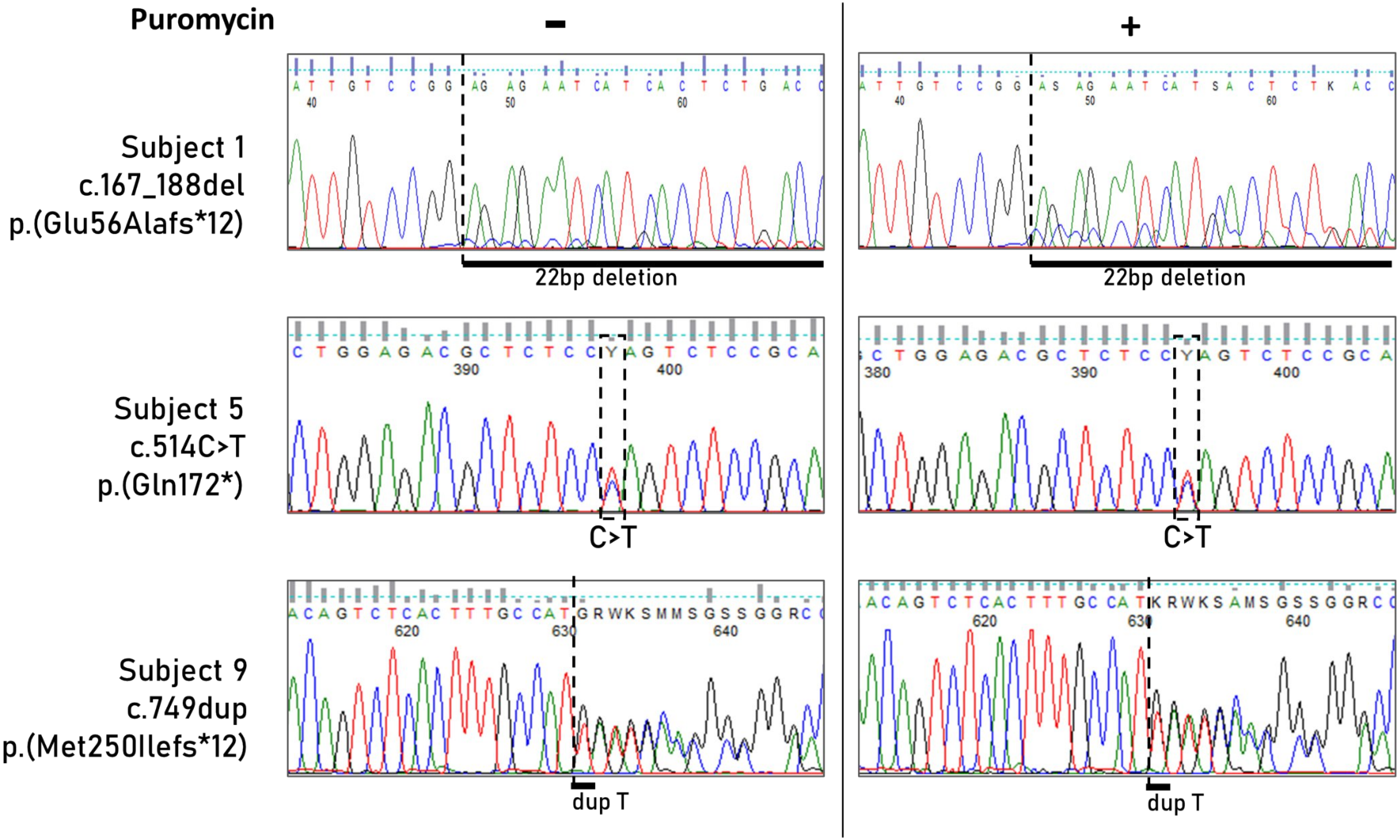

**Figure S3. Effect of puromycin treatment on variant *PCBP1* transcripts in subject-derived T cells.**

Sanger sequencing chromatograms for subject 1, 5 and 9, analysed in the absence (**−**, left panels) or presence (**+**, right panels) of puromycin incubation (4 hours at 1 mg/mL). In subject 1 (c.167_188del; p.(Glu56Alafs12)), puromycin leads to an increase in double sequence corresponding to the 22 bp deletion, indicating stabilization of the mutant transcript. In contrast, subjects 5 (c.514C>T; p.(Gln172*)) and 9 (c.749dup; p.(Met250Ilefs*12)) show similar levels of wild-type and variant with or without puromycin, suggesting no major effect of nonsense-mediated decay inhibition on these transcripts.

### Table S1. Molecular characteristics of candidate *de novo* variants in *PCBP1*.

| Family | Subject | Genomic alteration, Ch2(GRCh38) NC_000002.12 | cDNA variant* | Protein sequence change | Inheritance | Mode of detection | Variant number | Variantg Annotation link (MobiDetails) | MetaDome Score (missense variants only) | CADDphred v1.6 (missense variants only) | Control database occurence (GnomAD v4.1) | ACMG criteria** |
|---|---|---|---|---|---|---|---|---|---|---|---|---|
| **1** | **Subject 1** | g.70087910_70087931del | c.167_188del | p.(Glu56Alafs*12) | *de novo* | *trio WES* | 1 | https://mobidetails.chu-montpellier.fr/api/variant/675556/browser/ | NA | NA | 0 | PVS1, PS2, PM2 |
| **2** | **Subject 2** | g.70088007dup | c.264dup | p.(Asn89Glnfs*31) | *de novo* | *trio WES* | 2 | https://mobidetails.chu-montpellier.fr/api/variant/370449/browser/ | NA | NA | 0 | PVS1, PS2, PM2 |
| **3** | **Subject 3** | g.70088205dup | c.462dup | p.(Val155Cysfs*59) | *de novo* | *trio WGS* | 3 | https://mobidetails.chu-montpellier.fr/api/variant/1274334/browser/ | NA | NA | 0 | PVS1, PS2, PM2 |
| **4** | **Subject 4** | g.70088222A>C | c.479A>C | p.(Lys160Thr) | *de novo* | *trio WES* | 4 | https://mobidetails.chu-montpellier.fr/api/variant/675696/browser/ | 0,08 - Highly intolerant | 24.5 | 0 | PS2, PM1, PM2, PP2 |
| **5** | **Subject 5** | g.70088257C>T | c.514C>T | p.(Gln172*) | *de novo* | *trio WES* | 5 | https://mobidetails.chu-montpellier.fr/api/variant/675698/browser/ | NA | NA | 0 | PVS1, PS2, PM2 |
| **6** | **Subject 6** | g.70088287C>T | c.544C>T | p.(Pro182Ser) | *de novo* | *trio WES* | 6 | https://mobidetails.chu-montpellier.fr/api/variant/675700/browser/ | 0,29 - Intolerant | 24.6 | 0 | PS2, PM2, PP2 |
| **7** | **Subject 7** | g.70088293C>T | c.550C>T | p.(Gln184*) | *de novo* | *trio WES* | 7 | https://mobidetails.chu-montpellier.fr/api/variant/675711/browser/ | NA | NA | 0 | PVS1, PS2, PM2 |
| **8** | **Subject 8** | g.70088368dup | c.625dup | p.(His209Profs*5) | *de novo* | *trio WES* | 8 | https://mobidetails.chu-montpellier.fr/api/variant/1202342/browser/ | NA | NA | 0 | PVS1, PS2, PM2 |
| **9** | **Subject 9** | g.70088492dup | c.749dup | p.(Met250Ilefs*12) | *de novo* | *trio WES* | 9 | https://mobidetails.chu-montpellier.fr/api/variant/1202339/browser/ | NA | NA | 0 | PVS1, PS2, PM2 |
| **10** | **Subject 10** | g.70088650A>G | c.907A>G | p.(Asn303Asp) | *de novo* | *trio WES* | 10 | https://mobidetails.chu-montpellier.fr/api/variant/675725/browser/ | 0,12 - Highly intolerant | 27,2 | 0 | PS2, PM1, PM2, PP2 |
| **11** | **Subject 11** | g.70088659del | c.916del | p.(Arg306Alafs*37) | *de novo* | *trio WES* | 11 | https://mobidetails.chu-montpellier.fr/api/variant/675726/browser/ | NA | NA | 0 | PVS1, PS2, PM2 |
| **12** | **Subject 12** | g.70088677del | c.934del | p.(Gln312Argfs*31) | *de novo* | *trio WGS* | 12 | https://mobidetails.chu-montpellier.fr/api/variant/777565/browser/ | NA | NA | 0 | PVS1, PS2, PM2 |
| **13** | **Subject 13** | g.70088683A>T | c.940A>T | p.(Lys314*) | *de novo (same allele)* | *trio WES* | 13 (MNV) | https://mobidetails.chu-montpellier.fr/api/variant/675728/browser/ | NA | NA | 0 | PVS1, PS2, PM2 |
| | | g.70088695_70088706del | c.952_963 | p.(Pro318_Gly321del) | | | | https://mobidetails.chu-montpellier.fr/api/variant/675731/browser/ | NA | NA | 0 | PS2, PM2, PM4, PP3 |

a. ACMG: The American College of Medical Genetics and Genomics; NA: Not Applicable.
b.*Based on MANE Select/RefSeq transcript NM_006196.4
c.**PVS1 criteria based on predicted loss of function intolerance in GnomAD(v4)=1.
The criteria application and variant classification presented in this paper does not necessarily reflect the criteria and classification of the clinical testing laboratories involved in this paper
WES: whole exome sequencing; WGS: whole genome sequencing
